# Antidepressant drugs have pharmacological- and time-dependent effects on reinforcement learning in healthy volunteers: An 8 weeks randomized double-blind placebo-controlled study

**DOI:** 10.64898/2025.12.30.25342896

**Authors:** Lucie Berkovitch, Fabien Vinckier, Alexandre Salvador, Mathias Pessiglione, John F. W. Deakin, Gerard R. Dawson, Catherine J. Harmer, Guy M. Goodwin, Raphaël Gaillard

## Abstract

Isolating specific cognitive effects of antidepressant drugs is crucial to develop targeted and individualized treatment selection in psychiatry. In this double-blind, placebo-controlled study in healthy controls, we used computational modeling to characterize the cognitive effects of two classes of drugs for depression, escitalopram, a typical SSRI which increases serotonergic transmission, and agomelatine, which activates melatonin receptors and antagonizes 5-HT_2C_ serotonergic receptors. 128 healthy participants were randomized to receive either escitalopram (20 mg), agomelatine (25 mg or 50 mg) or placebo for 8 weeks and performed two complementary learning tasks at three time-points allowing to measure early (3 days), intermediate (2 weeks) and delayed (8 weeks) treatment effects. The first task was a simple probabilistic instrumental learning task evaluating how participants learned from positive and negative feedback. The second task was a more complex reversal learning task devised to assess learning from positive and negative feedback in an unstable environment. At 8 weeks, both drugs improved accuracy in task 1 and decreased choice stochasticity in task 2 compared to placebo. Agomelatine 25 and 50 mg had an additional early beneficial effect on reward processing at 3 days whereas agomelatine 50 mg showed maximal effects at 2 weeks. Our study provides one of the very first cognitive evaluations of the delayed effects of antidepressant drugs in healthy volunteers. It reveals that they share common beneficial effect on learning along with pharmacological-specific effects. All observed effects varied highly over time, highlighting the non-linearity of the cognitive impact.

## Introduction

Isolating specific cognitive effects of antidepressant drugs is crucial to develop targeted and individualized treatment selection in psychiatry. However, the mechanisms by which existing antidepressant drugs impact neurocognitive functions and how these effects correlate with their efficacy remain largely unknown [1, 2]. Computational modeling allows quantitative characterization of cognitive functions. It has already been used to link depressive symptoms and cognition [3, 4]. In particular, motivational deficits and anhedonia, which are hallmarks of depression, have been explored with reinforcement learning (RL) tasks, to distinguish between reward and punishment sensitivity [3, 5–8]. Such an approach demonstrated that patients with depression exhibit a decreased sensitivity to reward in relation to anhedonia [5, 9–14]. Additionally, it has been suggested that depression increases sensitivity to negative information [15, 16].

Studies investigating RL offer promising avenues to understand antidepressant mechanisms of action through the impact on cognition [17]. However, so far results have been mixed across clinical conditions. Indeed, some studies have found that SSRIs increased sensitivity to reward in various neuropsychiatric diseases [18–22], an effect that is also observed for drugs targeting noradrenaline [23, 24]. In contrast, other studies have suggested that SSRIs could decrease reward sensitivity while increasing sensitivity to punishment [25]. This last finding is consistent with observed antidepressant side effects such as apathy, emotional blunting and anhedonia [26–28]. Contradictory results regarding antidepressant effects on RL can be related to several factors: antidepressant drugs may show (i) different effects according to patients’ clinical conditions and clinical profiles (ii) effects that may vary over time, (iii) pharmacologically specific effects beyond generic antidepressant action. By generic action we mean those therapeutic effects that all antidepressant drugs may have in common.

To capture a more general pattern of antidepressants action and overcome variability across clinical conditions, several studies have examined antidepressant drug effects in healthy participants. Chamberlain and colleagues [29] demonstrated that acute inhibition of central serotonin reuptake impaired probabilistic learning in healthy humans. Other studies found that SSRIs decreased neural processing of aversive, rewarding and risk-associated stimuli [30–32]. Similar results were observed with tryptophan depletion [33] and selective serotonin releasing agents [34], whereas 5-HT_2A_ serotonin agonists such as LSD increase reward and punishment learning rates [35].

Antidepressant drugs likely have differential effects as a function of treatment duration. For instance, a study found that acute administration of a SSRI had no significant effect on RL in healthy participants, whereas one week of treatment enhanced learning from punishment and reduces learning from reward [36]. Other studies revealed that two weeks of treatment increased reward and effort learning signals [20], and three weeks of treatment decreased sensitivity to reinforcement without modulating reward or punishment learning rates [37]. Importantly, most studies had a single time-point of measure preventing evaluation of time-dependent effects in a unique within-subjects paradigm. Similarly, drug-induced manipulation of dopaminergic pathways modulates reward processing during RL [38–41], with opposite patterns observed acutely and at 2 weeks [42–44]. Moreover, long-term effects are usually assessed after 3 weeks of treatment [30, 31, 37], and to our knowledge, only one study explored delayed antidepressant effects in healthy volunteers more than 6 weeks after initiation [45]. Finally, because antidepressant drugs may have both generic and pharmacologically specific effects, comparing different drugs potentially allows these components to be distinguished. Indeed, direct comparisons suggest that noradrenergic and serotonergic drugs have distinct short-term effects on probabilistic learning and response to reward in healthy volunteers [29, 32].

To explore both time-dependent and pharmacological-specific effects on learning, we designed a double-blind, placebo-controlled study in healthy volunteers comparing drugs from two antidepressant classes at three different time-points to assess early (3 days), intermediate (2 weeks) and delayed (8 weeks) effects. Participants received escitalopram (a typical SSRI), agomelatine (25 or 50 mg) or placebo. Agomelatine does not act on the serotonin transporter but rather as a melatonin receptor agonist and a 5-HT_2C_ antagonist, resulting in an increase of noradrenaline and dopamine levels in the frontal cortex [for a review, see: , 46]. Interestingly, agomelatine has an early and selective effect on emotional processing in healthy volunteers [47]. In patients with depression, it is more efficacious than venlafaxine on pleasure and interest dimensions [48], including the sexual domain [49, 50]. Such effects may be related to its specific pharmacological mechanism of action, notably on the dopaminergic pathways.

In the current study, participants performed at each visit two distinct and complementary RL tasks assessing learning either in a stable or a changing environment. The first task was a simple probabilistic instrumental learning task devised to evaluate RL in a stable environment (contingency was fixed during the whole task for a given cue). This task, adapted from Pessiglione et al. [41], has already been extensively used to assess the ability to learn from positive and negative outcomes (gain/loss asymmetry) [19, 51] and has been demonstrated to be highly sensitive to dopamine modulation. The second task was a more complex reversal learning task devised to assess contingency learning in an unstable environment [52]. This task has previously been used to measure adaptation to uncertainty, as it is adaptive to adjust the weight of outcomes in value updating (learning rate) and the weight of value estimates in decision-making (choice stochasticity) in unstable environments, where contingencies are stochastic and susceptible to sudden reversals, depending on the confidence in value estimates. By doing so, we were able to disentangle early vs. delayed as well as generic vs. pharmacologic-specific effects of those drugs on learning from positive and negative feedback.

## Materials and Methods

### Participants and study design

Participants were recruited in two sites in England (Oxford and Manchester) through public advertisement. The study was approved by the local Ethics Committee and registered in the International Standard Randomized Controlled Trial Number Registry under the reference ISRCTN75872983. All participants gave their written informed consent prior to participation. Inclusion criteria were healthy volunteers, female or male, aged between 18 and 45 years, with a body mass index between 18 and 35 kg/m², non or moderate smokers (less than 10 cigarettes a day), fluent in English. They underwent a clinical examination including a structured clinical interview for DSM-IV-TR axis 1 disorder (SCID), a physical examination (heart rate, blood pressure), and biological samples (hematology, blood and urine biochemistry) to exclude any medical condition, psychiatric disease, use of psychotropic medication, drug abuse, or pregnancy (blood and urine pregnancy test). All women had to use effective contraception. No treatment was allowed during the study, except oral contraception, and paracetamol if required.

Following a double-blind placebo-controlled design, participants were randomized into four parallel arms to receive either agomelatine 25 mg, agomelatine 50 mg, escitalopram 20 mg (10 mg during the first week, 20 mg during weeks 2 to 8 and 10 mg during the last and 9th week) or placebo (see flow-chart in **Supplementary Figure 1**). Participants were assessed at three time-points while receiving treatments: 3 days (early), ∼2 weeks (intermediate, range: 12-18 days, mean = 15 days, SD = 2.2) and ∼8 weeks (delayed effects, range: 33-60 days, mean = 50.2 days, SD = 9.7) after treatment initiation. Participants performed another cognitive task assessing effort production [45]. Participants were evaluated for anxiety and depression levels using the Hospital Anxiety and Depression Scale (HADS) at baseline, one week and 8 weeks, and for sexual side effects using the Psychotropic-Related Sexual Dysfunction Questionnaire (PRSexDQ) at baseline, 2 weeks, 5 weeks and 8 weeks [50].

This study was exploratory and the sample size was estimated based on comparable studies [29, 47]. Exclusions and drop-outs before week 5 (discontinued intervention) were compensated to include 32 participants (16 females and 16 males) in each treatment arm, totaling 128 participants with completed intervention (i.e., who received at least 5 weeks of treatment, see **Supplementary Figure 1**). All those 128 participants were included in the analysis. One participant missed a visit at 3 days, and 4 participants missed a visit at 8 weeks.

At baseline, final groups did not significantly differ in either demographic characteristics, anxiety or depression levels (all *p* > 0.05, see **Table 1**).

**Table 1.** Participants’ demographic and psychometric characteristics at baseline. HAD = Hospital Anxiety and Depression scale; PRSexDQ = Psychotropic-Related Sexual Dysfunction Questionnaire

|  | Placebo<br>n = 32<sup>1</sup> | Escitalopram 20<br>n = 32<sup>1</sup> | Agomelatine 25<br>n = 32<sup>1</sup> | Agomelatine 50<br>n = 32<sup>1</sup> | p-value<sup>2</sup> |
| --- | --- | --- | --- | --- | --- |
| Sex (F/M) | 16/16 | 16/16 | 16/16 | 16/16 | 1 |
| Age mean | 23.0 (4.1) | 24.2 (4.1) | 24.2 (4.9) | 21.8 (3.8) | 0.09 |
| HAD<br>depression<br>score | 1.19 (1.35) | 0.84 (1.65) | 0.81 (1.15) | 0.91 (1.20) | 0.67 |
| HAD anxiety<br>score | 3.53 (2.55) | 3.78 (2.76) | 3.72 (2.53) | 3.50 (2.33) | 0.96 |
| PRSexDQ<br>score | 0.33 (0.61) | 0.80 (2.14) | 0.50 (0.88) | 0.29 (0.76) | 0.40 |

<sup>1</sup>n (%); Mean (sd)
<sup>2</sup> $\chi^2$ and ANOVA test

**Table 2.**
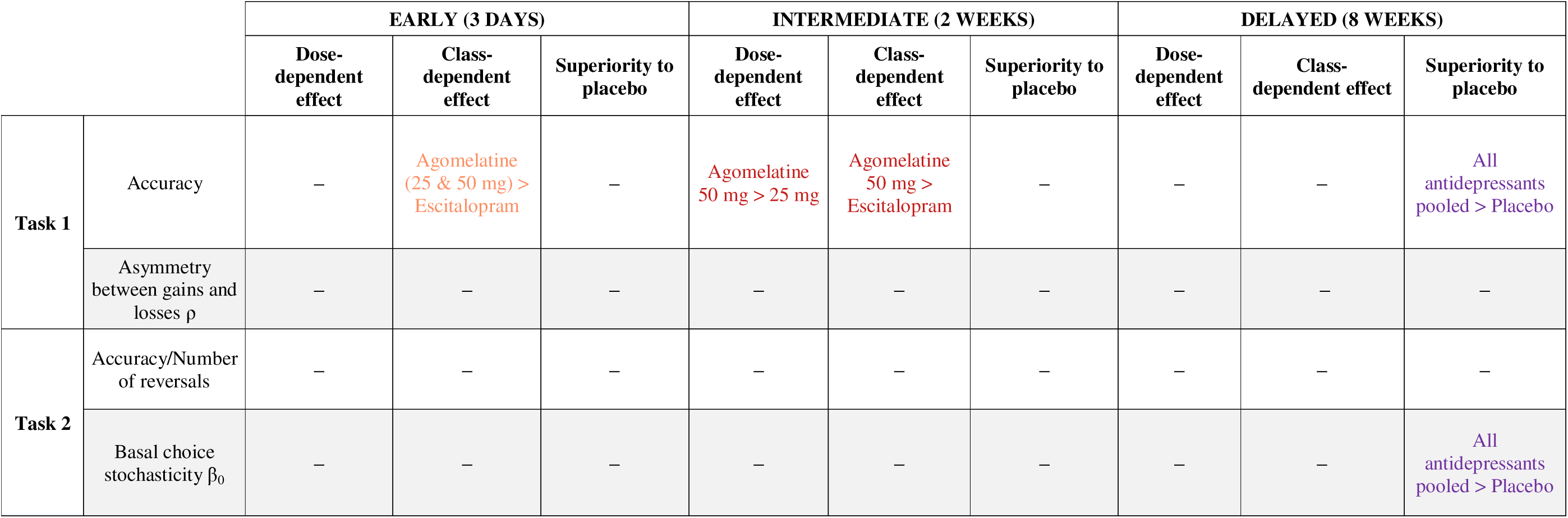
Summary of the results. . Colors represent the treatment which shows superiority (agomelatine 25 mg in orange, agomelatine 50 mg in red, all antidepressant in purple). “−” means that no significant effect of treatment was observed for this variable.

### Learning tasks

At each visit, participants performed two cognitive tasks evaluating learning abilities: a simple probabilistic instrumental learning task and a more complex reversal learning task (see **Figure 1**). Training was performed at each visit for each task before actual testing.

**Figure 1.**
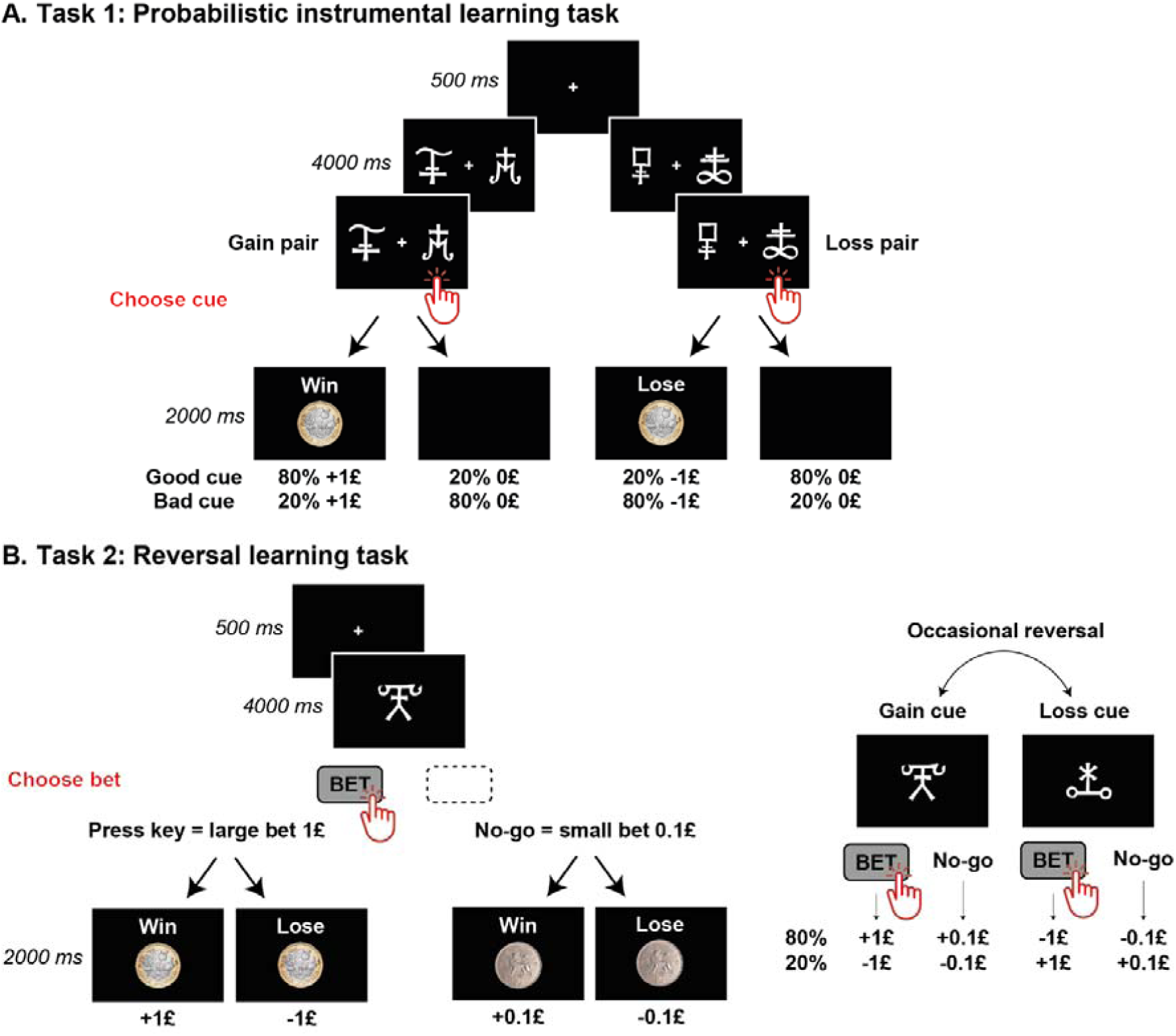
Learning tasks description |. **A.** Probabilistic instrumental learning task (task 1). In this task, participants have to choose between two visual cues associated with probabilistic gains or losses. One pair is associated with gain (+£1 or £0, gain pair) while the other pair is associated with loss (-£1 or £0, loss pair). For gain pairs, one cue is associated with an 80% chance of winning £1 (and £0 in 20% of the cases) whereas the other cue is mostly associated with a null outcome (£0 in 80% of the cases, £1 in 20% of the cases). Conversely, for loss pairs, one cue is associated with an 80% chance of losing £1 (and £0 in 20% of the cases) whereas the other cue is mostly associated with a null outcome (£0 in 80% of the cases, £1 loss in 20% of the cases). At each trial, a pair of cue is displayed on the screen and participants are asked to choose one of them. Participants have to learn, by trial and error, the cue-outcome associations in order to maximize their reward by choosing the cue associated with an 80% chance of winning in the gain pair, and the cue associated with only a 20% chance of losing in the losing one. **B.** Reversal learning task (task 2). In this task, participants have to choose how much to bet on a cue whose probabilistic outcome changes over time. One cue is associated with an 80% chance of winning (and 20% chance of losing, gain cue), the other cue has opposite contingencies (80% chance of losing, 20% chance of winning, loss cue). At each trial, participants have to decide whether they want to bet £1 (large bet) or £0.1 (small bet) on the displayed cue. Participants have to learn, by trial and error, the cue-outcome associations in order to maximize their outcome by placing large bets on the gain cue and try to win £1 and small bets on the loss cue to lose only £0.1. Unannounced contingency reversals (i.e. gain cue becoming loss cue and vice-versa) occurs after the subject responded accurately 14 to 17 times in a row, or at least every 60 trials.

#### Probabilistic instrumental learning task (Task 1)

To assess how participants learned from positive and negative feedback in a stable environment, they performed a probabilistic instrumental learning task where they had to choose between two visual cues associated with gain or loss [19, 41, 51]. Two pairs of abstract cues (letters from the Agathodaimon font) constituted gain and loss pairs respectively. For the gain pair, one cue was associated with an 80% chance of winning £1 (and 20% of null outcome, i.e. £0) whereas the other cue was mostly associated with a null outcome (£0 in 80% of the cases, £1 in 20% of the cases). Conversely, for the loss pair, one cue was associated with an 80% chance of losing £1 (and 20% of null outcome, £0) whereas the other cue was mostly associated with a null outcome (£0 in 80% of the cases, £1 loss in 20% of the cases). Participants had to learn, by trial and error, the cue-outcome associations in order to maximize their rewards by choosing the cue associated with an 80% chance of winning in the gain pair, and the cue associated with only a 20% chance of losing in the loss pair. At each visit, participants performed three blocks of the task in which new cues were presented. Each block included 60 trials (30 gain pair trials and 30 loss pair trials, intermixed in randomized order) totaling 180 trials per visit.

#### Reversal learning task (Task 2)

To probe participants’ ability to update decision-making according to positive and negative outcomes changing over time, they also performed a reversal learning task, where gain and loss associated with cues sequentially changed throughout the task [52]. One gain cue was associated with an 80% chance of winning (and a 20% chance of losing), whereas the other cue – loss cue – had opposite contingencies (i.e., 80% chance of losing, 20% chance of winning). In each trial, participants had to choose how much they wanted to bet on the cue displayed on screen (large/risky bet £1 or small bet £0.1). They had to learn, by trial and error, the cue-outcome associations in order to maximize their outcome by placing large bets on the gain cue and small bets on the loss cue. Unannounced contingency reversals, where the gain cue becomes the loss cue and vice-versa, occurred after the subject responded accurately 14 to 17 times in a row, or at least every 60 trials. Each participant performed 240 trials per visit, with in total at least 4 reversals.

#### Instructions

Participants were explicitly told that they would not receive the amounts earned during each task. Instead, they were paid a fixed amount that compensated for the time and expenses they devoted to the study. At each visit, before performing the actual task, participants were informed that cue-outcome relationships were not necessarily constant. However, they were not warned that contingencies could be reversed.

### Models

To obtain an assessment of treatment-specific effects on learning, we modeled participants’ behavior with reinforcement learning models based on individual sequences of choices according to outcomes.

#### Probabilistic instrumental learning task (Task 1)

For task 1, the model space consisted of 4 models. All models are variants of a basic, model-free reinforcement learning algorithm [53]. For each cue (say A or B) in each pair (say pair A-B or pair C-D), the model estimates the expected value, based on the individual outcome history, and makes a choice between choosing cue A or cue B when the pair was presented. The expected values are set at zero before learning, and after each trial t the value of the chosen cue (say A) is updated in proportion to prediction error, according to the “delta” (δ) rule [54, 55]:

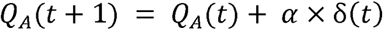

where δ(t) is the prediction error, defined as the difference between the actual and the expected outcome: δ(t) = R - Q_A_(t)

Then the probability, or likelihood, of choosing cue A when presented with the pair AB is estimated from the expected value according to the softmax rule:

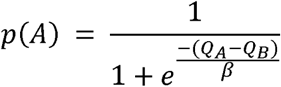

The learning rate α and the choice stochasticity β are free parameters, with the constraints 0 ≤ α ≤ 1 and β > 0. The learning rate adjusts the weight assigned to prediction error in value updating, and the choice stochasticity the degree of exploration (as opposed to exploitation of the learned value).

The four variants consisted in updating (or not) the value of the unchosen cue according to the update of the value of the chosen and to have a reinforcer equal to the financial outcome or representing gain/loss asymmetry in value updating (see **Supplementary Materials** for details)

#### Reversal learning task (Task 2)

The model space for Task 2 is the same as that used in a previous publication by Vinckier et al. [52]. This model is a hierarchical model with a reinforcement learning level and a meta-learning level. The basic, model-free reinforcement learning algorithm was similar to that used in Task 1, except that the choice was made between a riskier and a safer option rather than between two cues within a pair. We devised three variants of this reinforcement learning level, following step-by-step increments from model-free to model-based strategy, i.e. adding pieces of information about task structure. These variants included either the monetary value of the outcome (1, 0.1, -0.1 or -1), the outcome valence (Val) regardless of its magnitude (i.e. 1 when winning £1 or £0.1; -1 when losing £1 or £0.1) as reinforcer (see **Supplementary Materials** for details).

The meta-learning level was devised to account for subjects’ adaptation to volatility. Indeed, once subjects believe themselves to have a reasonably good estimation of contingencies, prediction errors should be tempered, and choices tuned to a more deterministic exploitation of learned contingencies [56–58]. Conversely, when contingencies suddenly change after reversals, prediction errors should be given more weight, and choices should be more exploratory. The meta-cognitive level consists in updating confidence so as to down-regulate contingency learning and choice stochasticity. We compared two ways to monitor confidence (according to prediction error or to outcome optimality) and four ways to use it (see **Supplementary Materials** for details). All in all, the model space contained 27 models.

#### Model comparison

All models were inverted using a variational Bayes approach under the Laplace approximation (Daunizeau et al., 2014; Friston et al., 2007; http://sites.google.com/site/jeandaunizeauswebsite/). This algorithm not only inverts nonlinear models but also estimates their evidence, which represents a trade-off between accuracy (goodness of fit) and complexity (degrees of freedom). The log-evidences estimated for each participant and model were submitted to a group-level random-effect analysis for each treatment group. This analysis was used to generate exceedance probability, which measures the plausibility that a given model (or model family) is more frequently implemented by participants that any other model (or model family) in the comparison set [61]. Fitted parameters (means of posterior distributions) from the best models were entered into random effect analyses (ANOVA and post-hoc *t*-tests) (see **Supplementary Materials** for details).

### Statistical analysis

This study was exploratory and the results presented here derive from post-hoc analyses (i.e., were not planned in the initial statistical analysis plan). Indeed, most computational tools used in the current study were developed after the validation of the analysis plan.

Analyses were conducted on 128 participants who had not discontinued at 5 weeks (per protocol set, see **Supplementary Figure 1**). Several participants did not properly perform the tasks: they had an average accuracy below 3 standard deviations of the mean of the rest of their group (i.e., for a given treatment arm, visit, pair valence). Their data in the corresponding condition were excluded. Exclusions represented less than 1.5 % of the total amount of data.

Repeated measure ANOVAs were performed separately for each task with accuracy (optimal response rates) and model parameters as dependent variables, treatment and visit, and valence as independent variables and age and sex as covariates. We first examined the full ANOVA where treatment was treated as a four-level factor variable (escitalopram vs. agomelatine 25 mg vs. agomelatine 50 mg vs. placebo). When a treatment × visit interaction was observed, we analyzed each visit separately using the following approach (see **Figure 2**):

1. First, we checked whether treatment effect was detected at a given visit.
2. Then, we assessed dose-related effects by specifically comparing agomelatine 25 mg and agomelatine 50 mg. If no significant difference was found, both doses were pooled.
3. We subsequently measured class-specific effects by comparing escitalopram and agomelatine, either pooling the two doses if no dose-specific effect was observed or treating them as separate groups if a dose-related effect was identified. If no difference was found, results from both drugs were pooled.
4. Finally, to estimate antidepressant effect over placebo, we compared each remaining active treatment arm to placebo, either pooling the three treatment arms together in the absence of significant class-effect (generic antidepressant effect) or as separate groups otherwise (class-specific effect).

**Figure 2.**
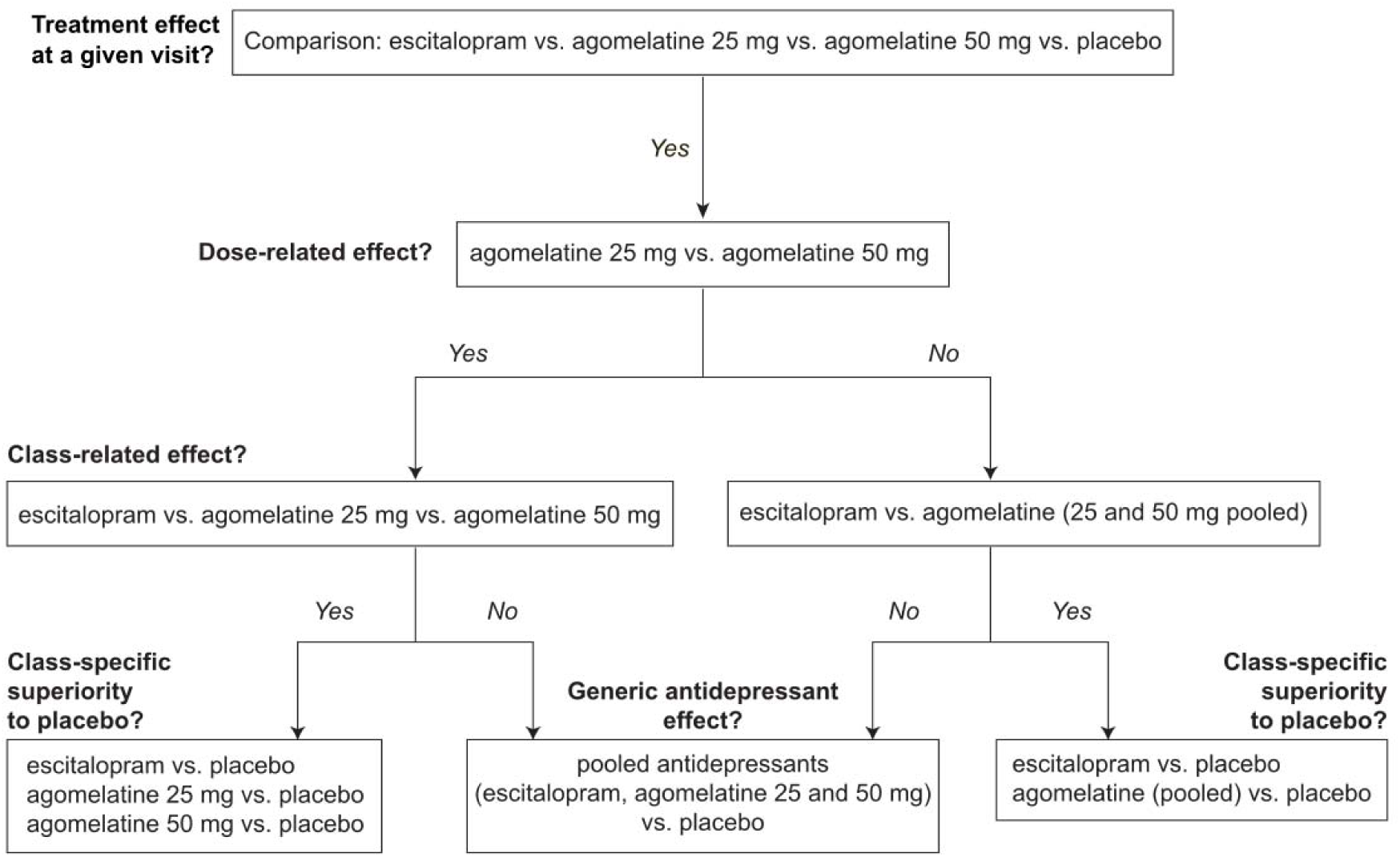
Four-step analysis pipeline. Statistical analyses were conducted following a systematic pipeline that estimated (1) main treatment effect, (2) dose-related effects, (3) class-related effects (pooling or not the two agomelatine doses depending on dose-related effects), (4) class-specific or generic antidepressant effect over placebo, pooling or not antidepressant classes together depending on class-related effects.

Additionally, we examined clinical variations assessed by HAD anxiety and depression scores and sexual side effects.

## Results

### Probabilistic instrumental learning task (Task 1)

#### Accuracy shows time-related and pharmacological effects which are dose and class-dependent

We first explored how participants’ accuracy varied as a function of treatment group (escitalopram vs. agomelatine 25 mg vs. agomelatine 50 mg vs. placebo), visit and valence (gain versus loss pairs). We observed a main effect of visit, indicating that participants improved with time (*F*_2,231_ = 15.19, *p* < 0.001), a main effect of valence whereby participants were better at picking the best cue in gain pairs compared to loss pairs (*F*_1,116_ = 15.27, *p* < 0.001) and a main effect of treatment (*F*_3,108_ = 2.70, *p* = 0.049). Importantly, there was a significant interaction between treatment and visit (*F*_6,231_ = 2.89, *p* = 0.010) and visit and valence (*F*_6,228_ = 3.54, *p* = 0.031), while the interaction between treatment and valence and the triple interaction between treatment, visit and valence did not reach significance (treatment × valence: *F*_3,116_ = 0.67, *p* = 0.57; triple interaction: *F*_6,228_ = 1.87, *p* = 0.088, see **Figure 3**). We then explored the effects of treatment in interaction with valence at each visit separately.

**Figure 3.**
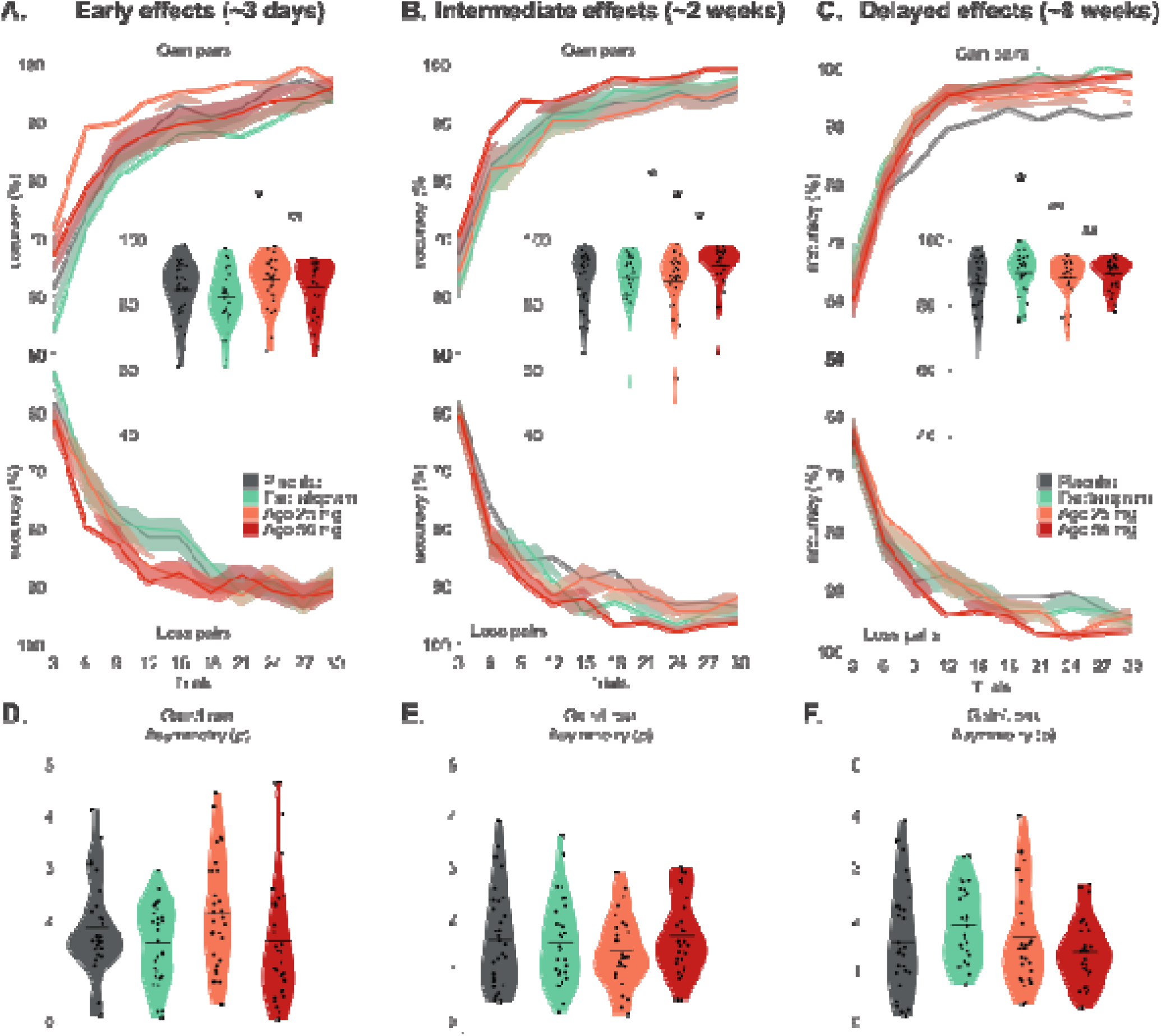
Treatment effects in the simple probabilistic learning task |. **A.** Accuracy at 3 days for the four groups (placebo in gray, escitalopram in green, agomelatine 25 mg in orange, agomelatine 50 in red). Learning curves depict the probability of making an optimal choice i.e. to choose the cue associated to an 80% chance of winning for the gain pair (top), or the cue associated to a 20% chance of losing in the loss pair (bottom), by bins of 3 trials, pooling the three blocks. Shaded areas represent inter-subject s.e.m. Insert: Violin plots summarize accuracy distribution averaged across valences. There was a significant class-dependent effect (agomelatine 25 and 50 mg pooled together vs. escitalopram). Each dot corresponds to a participant. Black horizontal bars represent means for each group. Stars represent significant statistical results (*p* < 0.05). **B. C.** Accuracy at 2 and 8 weeks for the four groups. Same description as for A. At 2 weeks there was both a dose-dependent (agomelatine 25 mg vs. 50 mg) and a class dependent effect (agomelatine 50 mg. vs. escitalopram; and agomelatine 50 mg vs. placebo). At 8 weeks, there was generic antidepressant effects (all active arms pooled together vs. placebo). **D.** Distribution of gain/loss asymmetry parameters by groups, at 3 days. The best model for task 1 was a Q-learning algorithm with three free parameters: the asymmetry between gains and losses ρ, the choice stochasticity β, and the learning rate α (not shown here because there was no significant difference between groups, see **Supplementary** for details). Violin plots summarize parameters’ distribution. Each dot corresponds to a participant. Black horizontal bars represent means for each group. **E. F.** Distribution of gain/loss asymmetry parameters by groups, at 2 and 8 weeks. Same description as for D.

At 3 days, there was a significant effect of treatment (*F*_3,117_ = 2.99, *p* = 0.034). The effect of agomelatine dose (25 mg vs. 50 mg) was not significant (*p* > 0.1), therefore, the two doses were pooled. We found a significant class effect, characterized by higher accuracy with agomelatine (25 mg and 50 mg pooled) compared to escitalopram (*F*L,LL = 7.97, *p* = 0.006). However, neither direct comparison of agomelatine nor escitalopram with placebo reached significance (all *p* > 0.1).

At 2 weeks, there was a significant effect of treatment (*F*_3,118_ = 2.84, *p* = 0.041). Accuracy was higher with agomelatine 50 mg compared to agomelatine 25 mg (*F*□,□□ = 6.48, *p* = 0.014). The comparison among the three treatment groups revealed a significant class-specific effect (*F*□,□□ = 3.88, *p* = 0.024), driven by the effect of agomelatine 50 mg. Accuracy with agomelatine 50 mg was not only higher than with agomelatine 25 mg (dose-related effect) but also higher than with escitalopram (class-related effect: *F*□,□_8_ = 5.36, *p* = 0.024) and placebo (class-specific superiority effect: *F*□,_59_ = 6.42, *p* = 0.014). However, there was no difference between escitalopram or agomelatine 25 mg and placebo (*p* > 0.1).

At 8 weeks, the effect of treatment was marginal (F□,□□□ = 2.50, p = 0.063). The effect of agomelatine dose was not significant (*p* > 0.1); therefore, both doses were pooled. There was no significant class effect either (*p* > 0.1) However, a significant general antidepressant effect over placebo was observed when all active treatment arms were combined (F□,□□□ = 5.81, p = 0.018).

Treatment effects did not significantly interact with valence (all *p* > 0.1).

#### Asymmetry between gain and loss varies non-linearly with treatment and visit

To measure what learning components were involved in these differences of accuracy, we modeled participants’ behaviors during task 1. The best model was a reinforcement learning model (a Q-learning algorithm) with three free parameters: the learning rate α, the choice stochasticity β, and the asymmetry between gains and losses ρ. Specifically, the update-rule was:

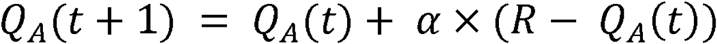

Where:

- α is the learning rate, comprised between 0 and 1
- R is the reinforcer. To account for an asymmetry between gains and losses, we used a free parameter ρ as the reinforcer instead of 1 when the outcome was strictly positive, while the monetary outcome (i.e. 0 or -1 respectively), was used when the outcome was negative or null.
- The value of the unchosen cue was updated using the counterfactual outcome, capturing the fact that participants realized that the two cues always had reciprocal contingencies, that is, information about the status of one cue also gave information about the other cue.

At each trial, the probability to choose A over B, *p*(A), was estimated using a softmax rule:

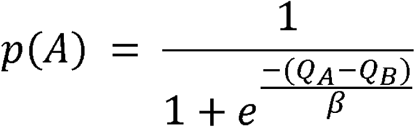

Where β is the temperature or choice stochasticity.

When β is high, p(A) is close to 0.5 regardless of the values of Q_A_ and Q_B_, implying random behavior. Conversely, when β is low, behavior is strongly driven by the difference in Q-values, meaning that regularities in previous trials are exploited to choose the best cue.

The only parameter significantly impacted by treatment was the asymmetry between gains and losses, ρ (see **Figure 3** and **Supplementary Table 1**), with a significant visit × treatment interaction (*F*_6,228_ = 2.48, *p* = 0.024). However, there was no significant main effect of treatment (*p* = 0.64), or visit (*F*_2,228_ = 2.83, *p* = 0.061), and the treatment effect did not reach significance when examined at each visit separately (all *p* > 0.1). Moreover, please note that this parameter accounts for valence effects, which did not significantly interact with treatments in the model-free analyses.

### Reversal learning task (Task 2)

#### Antidepressant action does not influence global performance

In task 2, accuracy was compensated along the task by reversals so both the accuracy and the number of reversals reflect performance. Both global accuracy and the number of reversals increase across visits (all *p* < 0.001) indicating an improvement of performance with time with time. However, there was no significant main effect of treatment or interaction between treatment and visits (all *p* > 0.1).

#### Antidepressant action decreases basal choice stochasticity after 8 weeks

The best model for task 2 was the same as in Vinckier et al. [52] (see **Supplementary Materials** and Vinckier et al. [52] for details). In this model, the valence of each cue is learned in a first layer of reinforcement learning with outcome valence as a reinforcer (1 for winning £1 or £0.1, -1 otherwise), and cue values are updated according to the outcome. Actual learning rate α and choice stochasticity β were not constant throughout the experiment as in the first task, as they were modulated by an estimate of the confidence C in the cue values. Confidence was updated using outcome optimality as a reinforcer (1 for winning £1 or losing only £0.1, 0 otherwise) but with a specific learning rate γ, and an initial confidence level C_0_. Confidence impact on the first-level free parameters (e.g. the learning rate α, and the choice stochasticity or temperature β) was controlled by a single weight κ. This model has therefore five free parameters: α_0_ (basal learning rate, corresponding to the learning rate when confidence = 0), β_0_ (basal choice stochasticity, corresponding to the choice stochasticity when confidence = 0), C_0_, γ, and κ.

The only parameter which was significantly impacted by treatment was the basal choice stochasticity β_0_ (see **Figure 4**). There was no main effect of treatment (*p* > 0.1), but a main effect of visit (*F*_2,236_ = 4.67, *p* = 0.010) and a significant interaction between treatment and visit (*F*_6,236_ = 2.27, *p* = 0.038). Given the significant interaction between treatment and visit, we explored the effects of treatment at each visit separately.

**Figure 4.**
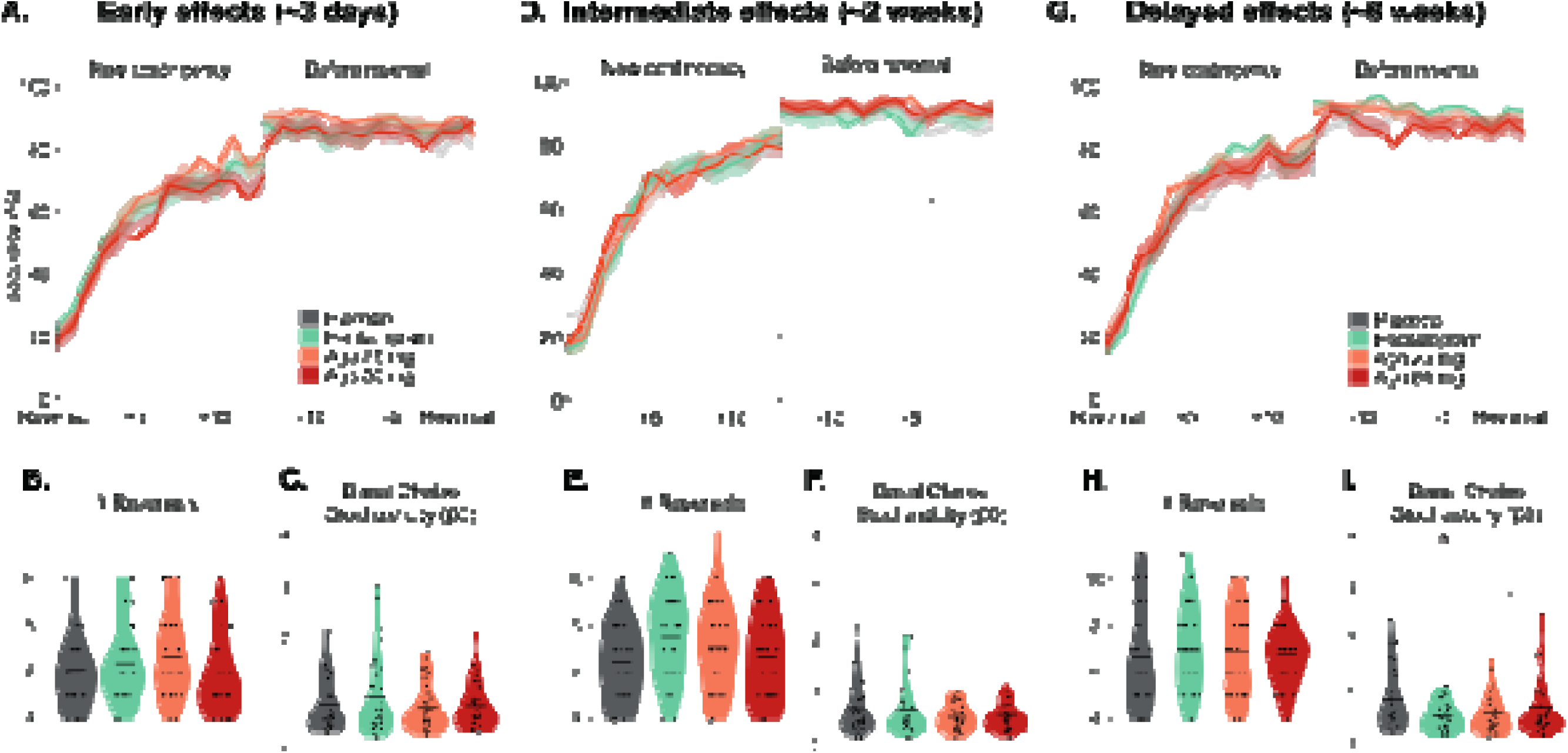
Treatment effects in the reversal learning task. | **A.** Performances at 3 days for the four groups (placebo in gray, escitalopram in green, agomelatine 25 mg in orange and agomelatine 25 mg in red). Learning curves depict the probability of making an optimal choice (a large bet on the gain cue or a small bet on the loss cue) in the 14 trials following the last reversal (*new contingency*, left part) or the 14 trials preceding next reversal (*before reversal*, right part). Please note that it is not possible to plot the whole curve as the length of the block varies as a function of accuracy. Shaded areas represent inter-subject s.e.m. **B.** Violin plot shows the number of reversals. Each dot corresponds to a participant. Black horizontal bars represent means for each group. **C.** Distribution of basal choice stochasticity parameters by groups, at 3 days. The best model in the reversal learning task was a hierarchical model with a reinforcement learning layer in which actual learning rate and choice stochasticity were not constant throughout the experiment but were modulated by an estimate of the confidence in the cue-values. It has five free parameters: basal learning rate (α_0_), basal choice stochasticity (β_0_), confidence learning rate (γ), the initial confidence level C_0_, and the weight of confidence on first layer parameters k. Parameters others than choice stochasticity are not shown here because there was no significant difference between groups (see **Supplementary Materials** for details). Violin plots summarize parameters’ distribution. Each dot corresponds to a participant. Black horizontal bars represent means for each group. **D. E.** Performances at 2 weeks for the four groups. Same description as for A. **F.** Model free parameters by groups, at 2 weeks. Same description as for C. **G. H.** Performances at 8 weeks for the four groups. Same description as for A. **I.** Model free parameters by groups, at 8 weeks. Same description as for C. Stars represent significant statistical results (*p* < 0.05). At 8 weeks, there was a significant generic antidepressant effect (all active arms pooled vs. placebo) on the basal choice stochasticity.

At 3 days and 2 weeks, there was no effect of treatment (both *p* > 0.1).

Conversely, at 8 weeks, there was a significant effect of treatment (*F*_3,115_ = 2.72, *p* = 0.048). The effect of agomelatine dose was not significant (*p* > 0.1), therefore, both doses were pooled. There was no significant class effect either (*p* > 0.1). However, a significant overall decrease in basal stochasticity was observed in comparing the combined antidepressant arms with placebo (*F*_1,117_ = 5.88, *p* = 0.017).

### Clinical variations and side effects

Anxiety and depression levels were assessed by the HAD at baseline (before treatment initiation), one week and 8 weeks. As expected in a healthy population, anxiety and depression scores were low (significantly lower than 7 in all groups at all visits, all *p* < 0.001). There were no significant changes of HAD depression scores with time (*F*_2,239_ = 1.01, *p* = 0.36). However, there were variations in the HAD anxiety scores across visits (*F*_2,240_ = 9.35, *p* < 0.001) with a marginal interaction between visit and treatment (*F*_6,240_ = 2.10, *p* = 0.055). This result was driven by a significant reduction of the HAD anxiety scores over time under escitalopram (*F*_2,60_ = 10.63, *p* < 0.001) and under agomelatine 50 mg (*F*_2,62_ = 3.42, *p* = 0.039) but not for the two other groups (all *p* > 0.3).

Sexual side effects were measured with the PRSexDQ at baseline (before treatment initiation), 2 weeks, 5 weeks and 8 weeks. As demonstrated in a previous publication on the same dataset, escitalopram yielded more sexual side effects than agomelatine regardless of the doses [50].

To explore the relationships between reward sensitivity and clinical evolution on the one hand and sexual side effects on the other hand, we correlated learning measures to HAD scores and to PRSexDQ scores whenever they were simultaneously measured, i.e., at 2 and 8 weeks for PRSexDQ and at 8 weeks for HAD scores. No significant direct correlation between learning measures and clinical scores simultaneously assessed survived correction for multiple comparisons.

There was no significant difference in terms of severe adverse events across treatment groups. Safety profiles of agomelatine 25-50 mg and escitalopram 20 mg were in line with their respective summary of product characteristics.

## Discussion

In this study, we examined the early (3 days), intermediate (∼2 weeks) and delayed (∼8 weeks) effects of escitalopram, agomelatine 25 mg, agomelatine 50 mg, and placebo on learning in healthy controls. We found that the drugs had significant effects on learning abilities with a complex pattern including both dose-dependent, class-related and generic antidepressant effects that vary over time. Agomelatine had early beneficial effects on probabilistic learning (task 1) compared to escitalopram. Conversely, at 2 weeks, we observed both a class-specific and a dose-dependent effects: agomelatine 50 mg improved performance compared to escitalopram and to agomelatine 25 mg. Finally, at 8 weeks, there was a generic antidepressant effect with no difference between antidepressant arms which showed a beneficial effect over placebo both for accuracy (task 1) and basal choice stochasticity (task 2). Importantly, there was no differential side effects, no variation in mood after several weeks of antidepressant treatment and no significant correlations between learning measures and clinical scores (depression, anxiety, sexual side effects).

Contrary to previous findings in healthy volunteers [29–32, 37], we did not observe a significant impairment in learning under SSRI in our study. Similarly, antidepressant effects did not significantly differ between reward and punishment processing. At most, there was a significant interaction between visit and treatment for asymmetry between gains and losses in task 1 and an interaction between treatment, visit and valence for global accuracy in task 2 but these results were not significant when examining each visit separately. Such findings are at odds with previous data and theoretical frameworks suggesting that antidepressant action rest upon a restoration of sensitivity to reward or positive events, favoring learning from positive events [7, 20, 62, 63].

Instead, learning was improved between 3 days and 2 weeks under agomelatine, and after 8 weeks under escitalopram. Indeed, despite totally different pharmacological mechanisms of action, agomelatine and escitalopram improved accuracy at 8 weeks in task 1 and decreased basal choice stochasticity in task 2 which captures a reduction of decision-making noisiness, and thus a general pro-cognitive effect. By contrast, other effects were dose-, class- and time-dependent. Specifically, agomelatine improved accuracy more than escitalopram at 3 days and 2 weeks, which could reflect both a positive effect of agomelatine or an initial negative effect of escitalopram on learning since neither were significantly different from placebo. These results fit previous clinical reports of an early motivational effect of agomelatine compared to other antidepressant drugs in patients with depression [48] and acute or early detrimental effects of SSRI on learning and reward processing [29, 32]. Furthermore, escitalopram may have been less tolerated than agomelatine at initiation and could have induced SSRI-induced emotional blunting [64].

The finding that escitalopram and agomelatine have inconsistent effects on learning across visits differs from a previous publication on the same dataset showing a sustained reduction of effort cost along escitalopram administration [45]. This suggests that the motivational and pro-learning effects may have distinct time courses. Noteworthy, our findings fit quite well with clinical experience. Indeed, patients with depression often face non-linear clinical responses during the first weeks of administration, with short improvements and exacerbations preceding sustainable response. Additionally, it is well-known from a clinical point of view that not all dimensions of depression respond with the same kinetics [65].

An important limitation of our study is that all the analyses were post-hoc and therefore did not derive from specific predictions regarding agomelatine versus escitalopram effects. Accordingly, our findings are hypothesis generating and will require replication to be convincing. In addition, as a study of reward the tasks may have a limited capacity to fully engage participants emotionally, because gains and losses were symbolic rather than material. In real world situations, the effects may be predicted to be more developed.

Cognition and motivation are core clinical dimensions of depression associated with functional impairment and subjective well-being [66, 67]. By investigating the impact of antidepressant drugs on learning in healthy volunteers, our study revealed common beneficial effects as well as pharmacological-specific and time-dependent effects of escitalopram and agomelatine. Although antidepressants may have differential effects in healthy controls and in patients with depression, cognitive testing may help identifying specific deficits to inform individualized treatment selection in psychiatry.

## Data Availability

All data produced in the present study are available upon reasonable request to the authors

## Acknowledgments

We thank all the participants. LB thanks the Fondation Bettencourt-Schueller, the Philippe Foundation, the Foundation L’Oréal-Unesco and the National Institute of Mental Health (R01MH116038 and U01MH121766) for their support.

