## Supplementary Materials for "Antidepressant drugs have pharmacological- and time-dependent effects on reinforcement learning in healthy volunteers: An 8 weeks randomized double-blind placebo-controlled study"

### Supplementary Information: Antidepressants have pharmacological and time dependent effects on reinforcement learning in healthy volunteers

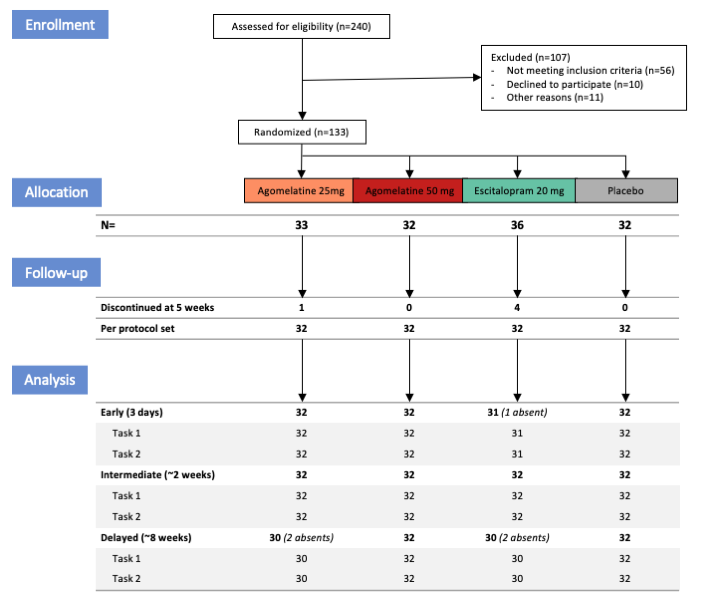

**Supplementary Figure 1. Flow-chart**. Participants were randomized into four parallel arms to receive either agomelatine 25 mg, agomelatine 50 mg, escitalopram 20 mg or placebo. Participants were assessed at three time-points while receiving treatments: 3 days (early), ~2 weeks (intermediate) and ~8 weeks (delayed effects) after treatment initiation. Exclusions and drop outs before week 5 (discontinued intervention) were compensated to include 32 participants completing intervention (16 females and 16 males) in each treatment group.

#### Probabilistic instrumental learning task (Task 1)

##### Task 1 model generation and selection

###### Description of the model space

The model space for task 1 includes 4 models. All models are variants of a basic, model-free reinforcement learning algorithm [1]. For each cue (say A or B) in each pair (say pair A-B or pair C-D), the model estimates the expected value, based on the individual outcome history, and makes a choice between choosing cue A or cue B when the pair was presented. The expected values are set at zero before learning, and after each trial t the value of the chosen cue (say A) is updated in proportion to prediction error, according to the “delta” (δ) rule [2, 3]:

$$Q_{A}(t+1) = Q_{A}(t)+ \alpha\times\delta\left( t \right)$$

where δ(t) is the prediction error, defined as the difference between the actual and the expected outcome: $\delta\left( t \right)= R- Q_{A}\left( t \right)$

Then the probability, or likelihood, of choosing cue A when presented with the pair AB is estimated from the expected value according to the softmax rule:

$$p\left( A \right) = \frac{1}{{1+e}^{\frac{-(Q_{A}-Q_{B})}{\beta}}}$$

The learning rate α and the choice stochasticity β are free parameters, with the constraints 0 ≤ α ≤ 1 and β > 0. The learning rate adjusts the weight assigned to prediction error in value updating, and the choice stochasticity the degree of exploration (as opposed to exploitation of the learned value).

We devised four variants on this basic Q-learning algorithm:

- The Q value of the unchosen symbol could either remained unchanged after the value of the chosen symbol has been updated, or it can also benefit from the information obtained on the chosen symbol, and be updated as well:

$$Q_{B}\left( t+1 \right)= Q_{B}\left( t \right)- \alpha\times\delta\left( t \right)$$

- The reinforcer R could either be equal to the financial outcome O of the choice, be it positive or negative. Alternatively, a new free parameter ρ could be used as the reinforcer instead of 1 when the outcome was strictly positive, while the monetary outcome (i.e. 0 or -1 respectively), was used when the outcome was negative or null. The free parameter ρ can be interpreted as a gain/loss asymmetry in value updating.

These two alternatives are used to create four different models in a 2 × 2 design.

###### Model Comparison

We first ensured that there was no difference for model distribution between treatment and visits [4, 5]. Therefore, we pulled all visits and treatments together in a random effect Bayesian model selection. The best model was the variant in which the unchosen symbol was updated as well as the chosen symbol, with a free parameter ρ to capture gain/loss asymmetry (xp = 0.98).

###### Parameter estimates for the best computational model.

|  |  | Learning rate (α) | | Choice stochasticity (β) | | Gain/Loss asymmetry (ρ) | |
| --- | --- | --- | --- | --- | --- | --- | --- |
| Visit | Treatment | Mean | Sd | Mean | Sd | Mean | Sd |
| Early | Agomelatine 25 mg | 0.13 | 0.08 | 0.45 | 0.31 | 2.05 | 1.06 |
|  | Agomelatine 50 mg | 0.14 | 0.06 | 0.41 | 0.26 | 1.51 | 1.25 |
|  | Escitalopram | 0.13 | 0.06 | 0.5 | 0.32 | 1.48 | 0.76 |
|  | Placebo | 0.12 | 0.06 | 0.52 | 0.42 | 1.78 | 0.91 |
| Intermediate | Agomelatine 25 mg | 0.13 | 0.04 | 0.35 | 0.18 | 1.32 | 0.68 |
|  | Agomelatine 50 mg | 0.12 | 0.04 | 0.26 | 0.1 | 1.6 | 0.73 |
|  | Escitalopram | 0.12 | 0.04 | 0.3 | 0.12 | 1.49 | 0.83 |
|  | Placebo | 0.13 | 0.07 | 0.41 | 0.33 | 1.53 | 0.94 |
| Delayed | Agomelatine 25 mg | 0.12 | 0.05 | 0.33 | 0.18 | 1.57 | 0.97 |
|  | Agomelatine 50 mg | 0.11 | 0.04 | 0.26 | 0.12 | 1.29 | 0.62 |
|  | Escitalopram | 0.11 | 0.03 | 0.34 | 0.23 | 1.82 | 0.72 |
|  | Placebo | 0.13 | 0.05 | 0.37 | 0.26 | 1.49 | 1.04 |

**Supplementary Table 1**. Parameter estimates for the best computational model in task 1.

#### Reversal learning task (Task 2)

##### Task 2 model generation and selection

###### Description of model space

The model space for Task 2 is the same as that used in a previous publication by Vinckier et al. [5]. This model is a hierarchical model with a reinforcement learning level and a meta-learning level.

###### Reinforcement learning level

We started with a basic, model-free reinforcement learning algorithm [1]. For each cue (say A or B), the model estimated the expected value, based on the individual outcome history, and made a choice between riskier and safer options. The expected values were set at zero before learning, and after each trial t the value of the ongoing cue (say A) was updated in proportion to prediction error, according to the “delta” (δ) rule [2, 3]:

$$Q_{A}(t+1) = Q_{A}(t)+ \alpha\times\delta\left( t \right)$$

where δ(t) is the prediction error, defined as the difference between the actual and the expected outcome: $\delta\left( t \right)= RQ(t)- Q_{A}\left( t \right)$

Then the probability, or likelihood, of larger bet was estimated from the expected value according to the softmax rule:

$$p\left( A \right) = \frac{1}{{1+e}^{\frac{-Q_{A}}{\beta}}}$$

The learning rate α and the choice stochasticity β are free parameters, with the constraints 0 ≤ α ≤ 1 and β > 0. The learning rate adjusts the weight assigned to prediction error in value updating, and the choice stochasticity the degree of exploration (as opposed to exploitation of the learned value).

We devised three variants of this reinforcement learning level, following step-by-step increments from model-free to model-based strategy, i.e. adding pieces of information about task structure.

In a first variant, the reinforcer RQ was the monetary value of the outcome (1, 0.1, -0.1 or -1). This variant can be considered as a model-free strategy, in line with the law of effect, meaning that outcomes increased the probability of repeating the same choice, depending on their sign and magnitude.

In a second variant, the reinforcer RQ was defined according to outcome valence (Val) and not its magnitude (i.e. 1 when winning £1 or £0.1 ; -1 when losing £1 or £0.1 ). This variant implies that subjects understood that cues determined the outcome valence (positive or negative) and not its magnitude, which depended on the choice.

In a third variant, the reinforcer RQ was defined according to outcome valence (Val) and the update of the current cue (say cue A) was transferred to the alternative cue (cue B):

$$Q_{B}(t+1) = {-Q}_{A}(t+1)$$

This variant implies that subjects understood that there were only two cues, with opposite valence. In other words, the two cue values summed up to zero.

###### Meta-learning level

Reinforcement learning models have constant parameters (learning rate and choice stochasticity). This limits the capacity to optimize the behavioral policy around the end of learning blocks, once subjects believe themselves to have a reasonably good estimation of contingencies. At this point, prediction errors should be tempered, and choices tuned to a more deterministic exploitation of learned contingencies [6–8].

Conversely, when contingencies suddenly change after reversals, prediction errors should be given more weight, and choices should be more exploratory. One way to optimize the behavior is to subordinate the reinforcement learning parameters to a higher level of control that monitors performance. A second series of models therefore included a meta-cognitive level consisting in updating confidence so as to down-regulate contingency learning and choice stochasticity. We compared two ways to monitor confidence and four ways to use it.

**Confidence monitoring level**

In both variants, confidence was monitored using a delta rule. The confidence learning rate γ was a free parameter, with 0 ≤ γ ≤ 1. The initial value of confidence, C_0_ was also fitted as a free parameter, with 0 ≤ C_0_ ≤ 1.

In a first variant, we used the absolute value of the prediction error computed at the reinforcement learning level to update confidence [9]:

$$C(t+1) = C(t)+ \gamma\times\left( \frac{\left( 2-\left| \delta\left( t \right) \right| \right)}{2}-C\left( t \right) \right)$$

In a second variant, we used outcome optimality (Op) to update confidence (i.e. 1 for winning £1 or losing £0.1 , -1 otherwise):

$$C(t+1) = C(t)+ \gamma\times\left( Op(t)-C\left( t \right) \right)$$

**Modulation of low-level free parameters**

Confidence was used to modulate the free parameters in the reinforcement learning models. This was done after each outcome, which brought information about how accurate the reinforcement learning model was, in terms of value estimates or behavioral policy. We considered four possibilities: modulation of learning rate or choice stochasticity, or both with the same weight, or both with a different weight.

The learning rate was modulated on the basis of not only confidence but also the outcome category. The idea is that to stabilize a representation of learned contingencies, subjects should increase their sensitivity to confirmation and decrease their sensitivity to contradiction. The impact of confidence on the learning rate 𝛼 therefore depended on whether the outcome was confirmatory (outcome and cue value have the same sign; $Val\left( t \right)=sign(Q\left( t \right))$) or not.

For confirmatory outcomes, 𝛼 was modulated as follows:

$\alpha_{m}\left( t \right)= \frac{\alpha_{0}+ k_{\alpha}\times C\left( t \right)}{1+k_{\alpha}\times C\left( t \right)}$where α_0_ and k_α_ are free parameters

And for contradictory outcomes:

$$\alpha_{m}\left( t \right)= \frac{\alpha_{0}}{1+k_{\alpha}\times C\left( t \right)}$$

Therefore, when confidence increased the modified learning rate 𝛼_m_ got closer to 1 for confirmatory outcomes and closer to zero for contradictory outcomes.

The choice stochasticity β was modulated such that exploration was reduced when confidence increased:

$\beta_{m}\left( t \right)= \frac{\beta_{0}}{1+k_{\beta}\times C\left( t \right)}$ where β_0_ and k_ß_ are free parameters

This modulation enables increasing exploitation above matching behavior, i.e. placing a larger bet more than 80% of the time following a cue that associated to a reward 80% of the time.

To test whether these modulations improved the fit of observed choices, we compared between models that included or not the free parameters (k_α_ and k_ß_), which could have or not identical values.

All in all, the model space contained 27 models.

###### Model Comparison

We first ensured that there was no difference for model distribution between treatment and visits [4, 5]. Therefore, we pulled all visits and treatments together in a random effect Bayesian model selection. The best model (xp = 1) was the variant in which the valence of each cue was learned in a first layer using outcome valence as the reinforcer (1 for winning £1 or £0.1, -1 otherwise), and updating both cue-values after outcome. In other terms, it takes into account the opposite valence of the two cues, the status of one cue informing about the status of the other. Actual learning rate and choice stochasticity were not constant throughout the experiment as in the first task, but were modulated by an estimate of the confidence in the cue-values. Confidence was updated using the outcome optimality as a reinforcer (1 for winning £1 or losing only £0.1, 0 otherwise) but with a different learning rate γ, and an initial confidence level C_0_. The impact of confidence on both first level free-parameters was controlled by a single weight κ.

###### Parameter estimates for the best computational model.

|  |  | Basal Learning rate (α_0_) | | Basal choice stochasticity (β_0_) | | Confidence learning rate (γ) | | Confidence weight (κ) | |
| --- | --- | --- | --- | --- | --- | --- | --- | --- | --- |
| Visit | Treatment | Mean | Sd | Mean | Sd | Mean | Sd | Mean | Sd |
| Early | Agomelatine 25 mg | 0.35 | 0.17 | 0.69 | 0.45 | 0.51 | 0.12 | 0.72 | 0.69 |
|  | Agomelatine 50 mg | 0.31 | 0.2 | 0.72 | 0.44 | 0.49 | 0.14 | 0.46 | 0.54 |
|  | Escitalopram | 0.4 | 0.16 | 0.88 | 0.76 | 0.52 | 0.13 | 0.65 | 0.71 |
|  | Placebo | 0.32 | 0.15 | 0.75 | 0.55 | 0.49 | 0.12 | 0.47 | 0.52 |
| Intermediate | Agomelatine 25 mg | 0.35 | 0.14 | 0.51 | 0.24 | 0.48 | 0.16 | 0.74 | 0.6 |
|  | Agomelatine 50 mg | 0.39 | 0.15 | 0.55 | 0.32 | 0.48 | 0.13 | 0.63 | 0.47 |
|  | Escitalopram | 0.4 | 0.14 | 0.68 | 0.53 | 0.5 | 0.11 | 0.68 | 0.58 |
|  | Placebo | 0.38 | 0.19 | 0.69 | 0.51 | 0.53 | 0.12 | 0.64 | 0.43 |
| Delayed | Agomelatine 25 mg | 0.39 | 0.17 | **0.57** | 0.38 | 0.52 | 0.13 | 0.62 | 0.57 |
|  | Agomelatine 50 mg | 0.34 | 0.14 | **0.69** | 0.58 | 0.49 | 0.15 | 0.49 | 0.48 |
|  | Escitalopram | 0.35 | 0.15 | **0.51** | 0.27 | 0.46 | 0.14 | 0.69 | 0.51 |
|  | Placebo | 0.41 | 0.15 | **0.83** | 0.57 | 0.48 | 0.15 | 0.74 | 0.57 |

**Supplementary Table 2.** Parameter estimates for the best computational model in Task 2. Parameter estimates for which there is a significant statistical difference between antidepressant treatments compared to placebo are in bold.
